# Evaluating Smartphone-Based Cough Monitoring for Tuberculosis Screening and Triage in Uganda: A Mixed-Methods Evaluation

**DOI:** 10.1101/2025.09.01.25334889

**Authors:** Patrick Biché, Francis Kayondo, Annet Nalutaaya, James Mukiibi, Mariam Nantale, Joowhan Sung, David W Dowdy, Achilles Katamba, Emily A Kendall

## Abstract

Symptom-based tuberculosis (TB) screening often relies on cough reporting, which may be unreliable. Passive cough monitoring via smartphone apps offers a less subjective alternative. This study evaluated the diagnostic accuracy and implementation feasibility of smartphone-based cough frequency monitoring for identifying individuals likely to have TB in Uganda. We enrolled adults (≥15 years) screened for TB in community settings or tested at health facilities in Kampala. Participants underwent microbiological testing and 48-hour cough monitoring using the Hyfe Research app. TB status was determined using Xpert MTB/RIF Ultra and culture, and inverse probability weighting was applied to adjust for differential enrollment. We compared cough frequency between individuals with and without TB and assessed diagnostic accuracy using weighted ROC curves. We also conducted staff interviews, analyzed thematically, to explore implementation challenges. Of 884 enrolled participants, 197 had valid cough recordings and TB status (101 from community screening, 96 from health facilities). Individuals with TB had higher median cough frequency than those without: 2.2 (interquartile range 0.8-6.1) vs 0.9 (0.4-2.0) coughs per hour in the community and 6.7 (2.6-27.5) vs 2.4 (0.9-4.8) in facilities; Wilcoxon P < 0.0001 for both. AUCs for smartphone-based cough recording were 0.69 (95% CI: 0.58–0.79) in the community and 0.76 (95% CI: 0.6–0.88) in health facilities. At 90% sensitivity, specificities were 20% (95% CI: 0–22%) and 46% (95% CI: 1–32%), respectively. Smartphone-recorded cough frequency was moderately correlated with self-reported cough severity, respiratory-related quality of life scores (Saint George’s Respiratory Questionnaire), and staff-observed coughs. Staff cited device visibility, stigma, and security concerns as barriers to implementation. While smartphone-recorded cough frequency was associated with TB status, it did not meet diagnostic accuracy thresholds for stand-alone screening or triage. Implementation challenges also limited data collection. Addressing operational barriers will be critical to future development and deployment of cough monitoring tools for TB screening.

## Introduction

Decisions about whether to test for tuberculosis (TB) are often guided by symptoms, particularly cough despite evidence that, during population-based screening, approximately half of individuals with microbiologically confirmed TB screen negative for symptoms [1,2]. However, cough may be unrecognized, unreported, or assessed inconsistently by different individuals [3]. This unreliability of subjective symptom reporting presents challenges for healthcare providers and TB screening programs. More objective cough measurement tools that improve symptom assessment may therefore enhance the performance of TB screening and triage strategies.

Tools to quantify cough have evolved over decades, progressing from mechanical counters and symptom diaries to digital applications that leverage artificial intelligence (AI) to identify, count, and characterize cough sounds [4]. Recent advancements in digital acoustic surveillance have further expanded these capabilities, with AI models now demonstrating promise in detecting disease-specific acoustic patterns [5]. The use of cough-based diagnostic tools expanded during the COVID-19 pandemic, reflecting growing interest in passive, digital respiratory surveillance [5]. The Hyfe Research App (“Hyfe”) is one such tool for which early studies demonstrated high accuracy in detecting cough sounds in a controlled setting [6–8]. However, the application of these tools for infectious disease screening, especially in real-world, resource-constrained settings, remains underexplored.

Digital cough measurement tools have potential to improve TB screening and triage in high-TB-burden, resource-limited settings where TB diagnostic capacity is constrained but digital access is increasing. However, evidence remains limited about their feasibility and diagnostic utility in these settings. This study evaluates whether cough frequency, measured via a smartphone-based passive monitoring app, can distinguish individuals with TB in both community-based screening and health facility-based triage settings, evaluating accuracy against an established target product profile (TPP) from the World Health Organization (WHO) [9]. We also interviewed staff to understand perceptions of cough recording and potential obstacles to its broader implementation.

## Methods

### Study population

This analysis includes individuals who participated in either community-based screening or routine clinic-based diagnosis for presumptive TB in the Kampala, Uganda, metropolitan area between February 2021 and December 2024. This study was embedded within parent studies that enrolled participants undergoing TB screening primarily in high-traffic areas within high-risk communities (the “community” cohort) and routine diagnostic evaluation in ambulatory health facilities (the “health facility” cohort). Both cohorts recruited individuals aged ≥15 years and used sputum Xpert® MTB/RIF Ultra (“Ultra”) testing as part of symptom-agnostic algorithms.

Both cohorts recruited all individuals with trace Ultra results during the enrollment period, as well as a selection of individuals with positive (greater than trace) or negative Ultra results. Individuals with negative results in the community setting, and with either positive (greater than trace) or negative results in health facilities, were matched by age, sex, and human immunodeficiency virus (HIV) status to those enrolled with trace-positive results. Enrolled participants from both cohorts were offered additional *M. tuberculosis* microbiological testing and smartphone app-based cough frequency measurement. Further details of the parent studies are described elsewhere [10,11].

### Data Collection

At the time of enrollment into both cohorts, participants underwent standardized interviews that included demographic information, symptom characterization, and TB treatment history.

Participants screened in the community after July 2021 were also asked, at the time of screening, whether they currently had a cough. Symptoms were characterized through a questionnaire about the presence and duration of individual symptoms as well as the Saint George’s Respiratory Questionnaire (SGRQ) [12] (designed to measure health-related quality of life in chronic pulmonary disease), a cough severity rating (self-assessed using a visual analogue scale from 0-100, set to zero for those not reporting cough [13]), and observations of cough by research staff during the enrollment (assessed as both a qualitative assessment and a five-minute count of coughs). The applicable subset of interview questions can be found in S1 Appendix. Additional testing included solid and liquid cultures, a repeat Ultra test on expectorated sputum, and a posteroanterior chest x-ray (CXR).

Participants were also asked to complete 48 hours of cough frequency recording following the enrollment visit. Consenting participants were provided with a study smartphone equipped with the Hyfe research app, a lapel microphone, and an external battery bank for charging. This app processes sound continuously and saves a <0.5 second recording whenever there is an “explosive” sound that exceeds background noise signal by a set threshold. Each of these sounds then undergoes AI-based analysis and is assigned a score between 0 and 1, correlated to the probability of being a cough. Participants were asked to keep the phone near them for 48 hours. They were also provided with a lavalier microphone, which they were asked to connect and wear whenever possible (to better differentiate sounds coming from near their own mouth) and a spare battery pack (for recharging if electricity was unavailable).

To better understand barriers to cough data collection, interviews were conducted with research staff who had used the Hyfe research app to measure cough frequency. Participating staff were asked about feedback they received from participants, factors they considered when deciding to give the device to participants, and questions related to the recording process and areas for improvement. Interviews were audio-recorded, transcribed verbatim, and analyzed using thematic analysis in NVivo software [14]. The interview guide is listed in S2 Appendix.

### TB status classification

In classifying participants’ TB status for diagnostic accuracy analyses, we considered Ultra and culture results. For participants with a history of TB treatment (for whom Ultra false-positive results are more likely), we considered culture results to be definitive [15]. Absent a history of TB treatment, those with a positive Ultra results greater than trace were classified as having TB, but those with an initial trace result required at least one additional trace result, or any other positive Ultra or culture result, to be classified as having TB. Those with a negative initial Ultra result and no prior treatment required another negative Ultra or culture result (and no positive results) to be classified as TB-negative and were excluded otherwise.

### Statistical Analysis

We applied a developer-recommended score cutoff of ≥0.85 to define a cough among device-captured explosive sounds. We limited analysis to time periods when the participant’s recording app was active, the device was documented by research staff as having been with the participant, and the participant did not report having been more than two meters away from the device. We then calculated a mean cough frequency in coughs per hour for each participant.

Because individuals with different results (negative, trace, positive) on a preceding Ultra test had different probabilities of recruitment into the parent study, analyses of diagnostic accuracy were weighted to reflect the original population that had undergone screening or diagnostic testing.

We determined the proportion of individuals with each Ultra result among all participants in our community-based screening intervention, and (separately) in a tally of 12 months of Ultra test results from patients evaluated for presumptive TB at five participating health facilities. We then compared these proportions to the proportion of each Ultra result type among participants who completed cough recording, and we weighted the data by this relative prevalence using inverse probability of selection weight. All diagnostic accuracy analyses were performed separately for community-based screening versus health facility-based diagnostic testing. Values for weighting can be found in S3 Table.

We compared cough frequency by TB status using weighted Wilcoxon rank sum tests. We also examined correlations between cough frequency and various subjective respiratory symptoms measures using Spearman correlation coefficients. We estimated the accuracy of cough frequency for classifying TB status by creating weighted receiver operating characteristic (ROC) curves and estimating areas under the curve (AUCs) with 95% confidence intervals (WeightedROC package version 2020.1.31) [16]. Sensitivity and specificity of subjective cough screening were calculated among participants with confirmed TB status who had undergone symptom screening at the time of initial Ultra testing in the community, with weighted binomial 95% confidence intervals. To compare screening accuracy between subjective cough reporting and measured cough frequency, we used ROC analysis to identify the cough frequency threshold that yield equivalent specificity to reported cough, and the sensitivity of that threshold was statistically compared to the sensitivity of reported cough using McNemar’s test for paired proportions. All statistical analyses were conducted using R version 4.5.0 [17].

### Ethical Considerations

Informed consent (or adolescent assent and parental informed consent) was obtained from all study participants. The study was approved by the Institutional Review Boards at the Johns Hopkins University School of Medicine and Makerere University School of Public Health.

## Results

### Completion of cough recording

A total of 400 community and 484 health facility participants were enrolled during the analysis period, of whom 147 (36%) community and 126 (26%) health facility participants received a device and completed at least one hour of cough frequency recording (Figure 1). No devices were available for six participants. When devices were available, the reason for not receiving a device was documented for 209 participants (documented only for the first eight months of the study): 110 (53%) were deemed ineligible by study staff, mainly due to safety concerns, and 99 (47%) declined by participants for reasons that included feeling unsafe carrying the device or objections of partners/family. After also excluding 76 participants with missing cough data or indeterminate TB status, 101 (25%) participants enrolled from community screening and 96 (20%) enrolled from health facility-based evaluation were included in this analysis. A total of 8625 hours of cough recording from these participants were analyzed, with a median recorded time of 45 hours (interquartile range [IQR] 27-49) per participant.

**Figure 1.**
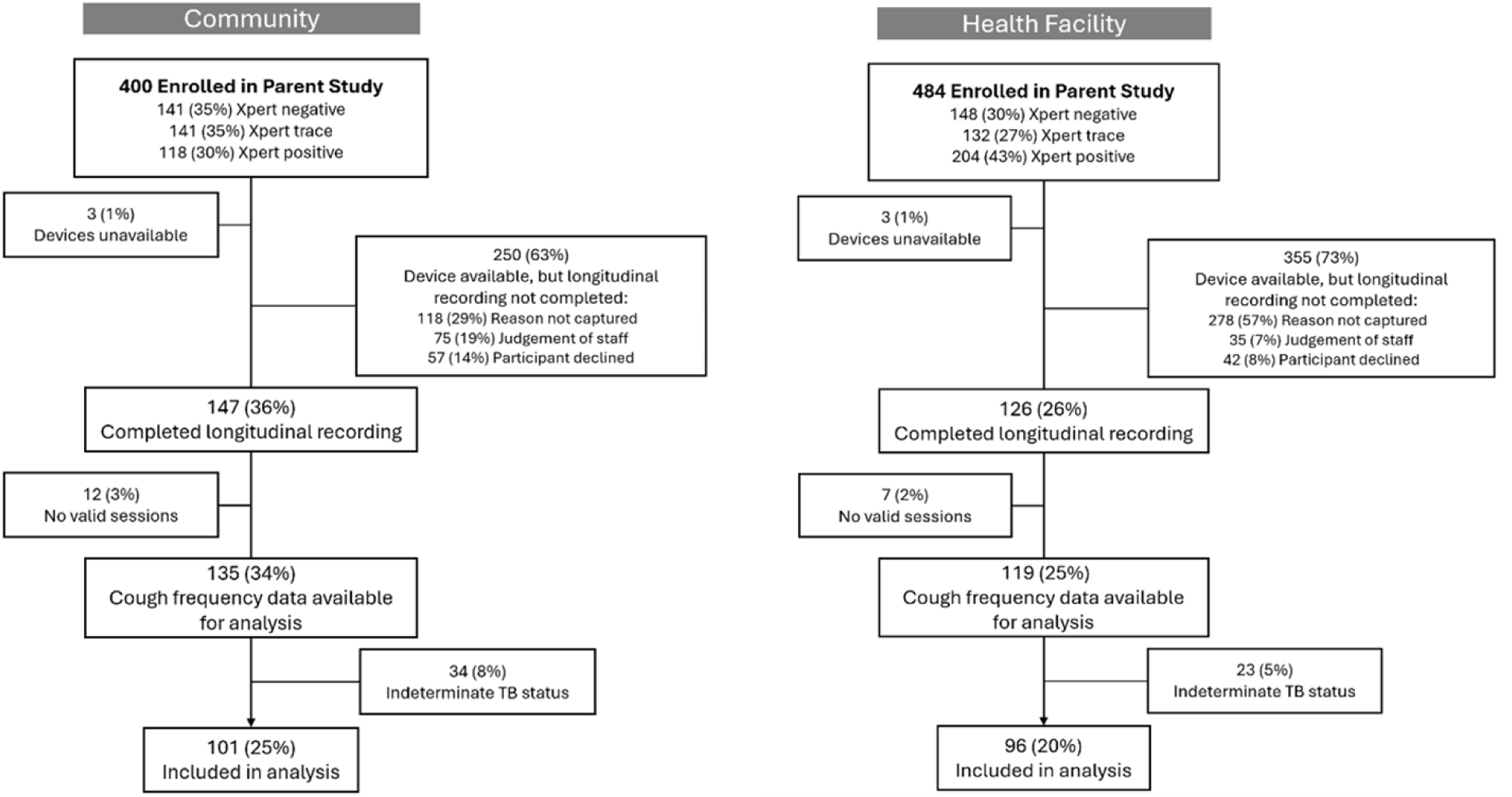
**Flowchart of study participants and reasons for exclusion from analysis.** Shown are the results of recruitment and enrollment into the present analysis of smartphone-based cough recording, relative to sampling in the parent study. “Reason not captured” for lack of completing longitudinal recording reflects the parent study design, in that only solicited cough sounds were recorded for some participants resulting in no documented reason for not completing longitudinal recording. No valid sessions occur when there are no recorded sessions that fall within the period research staff reported the participant had the device. Other reasons may include device malfunction, improper use of devices, or data loss.

### Participants

Included participants were 53% male with a median age of 32 (IQR 25-38) (Table 1). Overall, 27% were living with HIV (11% in the community cohort and 44% in the health facility cohort), 19% had a history of previous TB, and 21% reported current or former smoking. In the community cohort of 56 participants who were asked about symptoms at the time of screening, 24 (43%) reported having a cough. In the health facility cohort, cough was assessed only at enrollment, after initial Ultra test results, with 85% (82/96) of participants reporting cough.

**Table 1.** Characteristics of participants included in cough frequency analyses.

| Recruitment Source | Overall | Community |  | Health Facility |  |
| --- | --- | --- | --- | --- | --- |
| TB Status |  | Negative | Positive | Negative | Positive |
| N | 197 | 54 | 47 | 68 | 28 |
| Total hours recorded | 8625 | 2323 | 2281 | 2902 | 1119 |
| Median hours recorded (IQR) | 45 (27-49) | 45 (30-48) | 48 (27-53) | 44 (26-48) | 46 (33-50) |
| Male, n (%) | 104 (53) | 30 (56) | 31 (66) | 29 (43) | 14 (50) |
| Age, median (IQR) | 32 (25-38) | 28.5 (24-35) | 32 (24-37.5) | 35 (27-40.25) | 30 (23-36.5) |
| **HIV positive, n (%)** | 53 (27) | 7 (13) | 4 (9) | 32 (47) | 10 (36) |
| **Previous TB, n (%)** | 38 (19) | 7 (13) | 0 (0) | 29 (43) | 2 (7) |
| **Smoker, n (%)** | 41 (21) | 6 (11) | 15 (32) | 13 (19) | 7 (25) |
| **Reported cough\*, n (%)** | 106/152 (70%) | 12/29 (41) | 12/27 (44) | 55 (81) | 27 (96) |
| **Cough severity, median (IQR)** | 25 (8-6) | 0 (0-10) | 40 (20-60) | 32 (10-50) | 50 (30-70) |
| **Cough duration in weeks, median (IQR)** | 4 (3-10) | 2 (1-4) | 4 (2-12) | 3 (2-7) | 4 (2-10) |
| **SGRQ†, median (IQR)** | 15 (6-44) | 10 (6-20) | 22 (12-65) | 15 (6-49) | 34 (26-53) |
| **Ultra >trace, n (%)** | 49 (25) | 0 (0) | 28 (60) | 0 (0) | 21 (75) |
| **Ultra trace, n (%)** | 11 (6) | 0 (0) | 4 (9) | 3 (4) | 4 (14) |
| **Culture positive, n (%)** | 55/190 (29) | 0/52 (0) | 30/46 (65) | 0/64 (0) | 25/28 (89) |
| **Abnormal CXR‡, n (%)** | 91/185 (49) | 11/53 (21) | 36/45 (80) | 24/64 (38) | 20/23 (87) |

\* Reported during screening in the community cohort and during enrollment in the health facility cohort
† Saint George's Respiratory Questionnaire, , with higher scores (on a scale from 0 to XX) representing greater impairment in respiratory-related quality of life
‡ Chest x-ray

Among those from either cohort who reported cough at study enrollment, the median self-reported severity was 25 (IQR 8-60) on a scale of 0 to 100 and the median duration was 4 weeks (IQR 3-10). CXR was abnormal in 46% of participants.

### Cough frequency

In the community cohort, participants with TB had higher weighted median cough frequency on enrollment than TB-negative participants (2.2 coughs per hour (IQR 0.8-6.1) vs 0.9 coughs per hour (IQR 0.4-2.0), *P* < 0.0001; Figure 2a). A similar pattern held in the health facility cohort, with a weighted median of 6.7 (IQR 2.6-27.5) vs 2.4 (IQR 0.9-4.8) coughs per hour among participants with versus without TB (*P* < 0.0001; Figure 2b).

**Figure 2.**
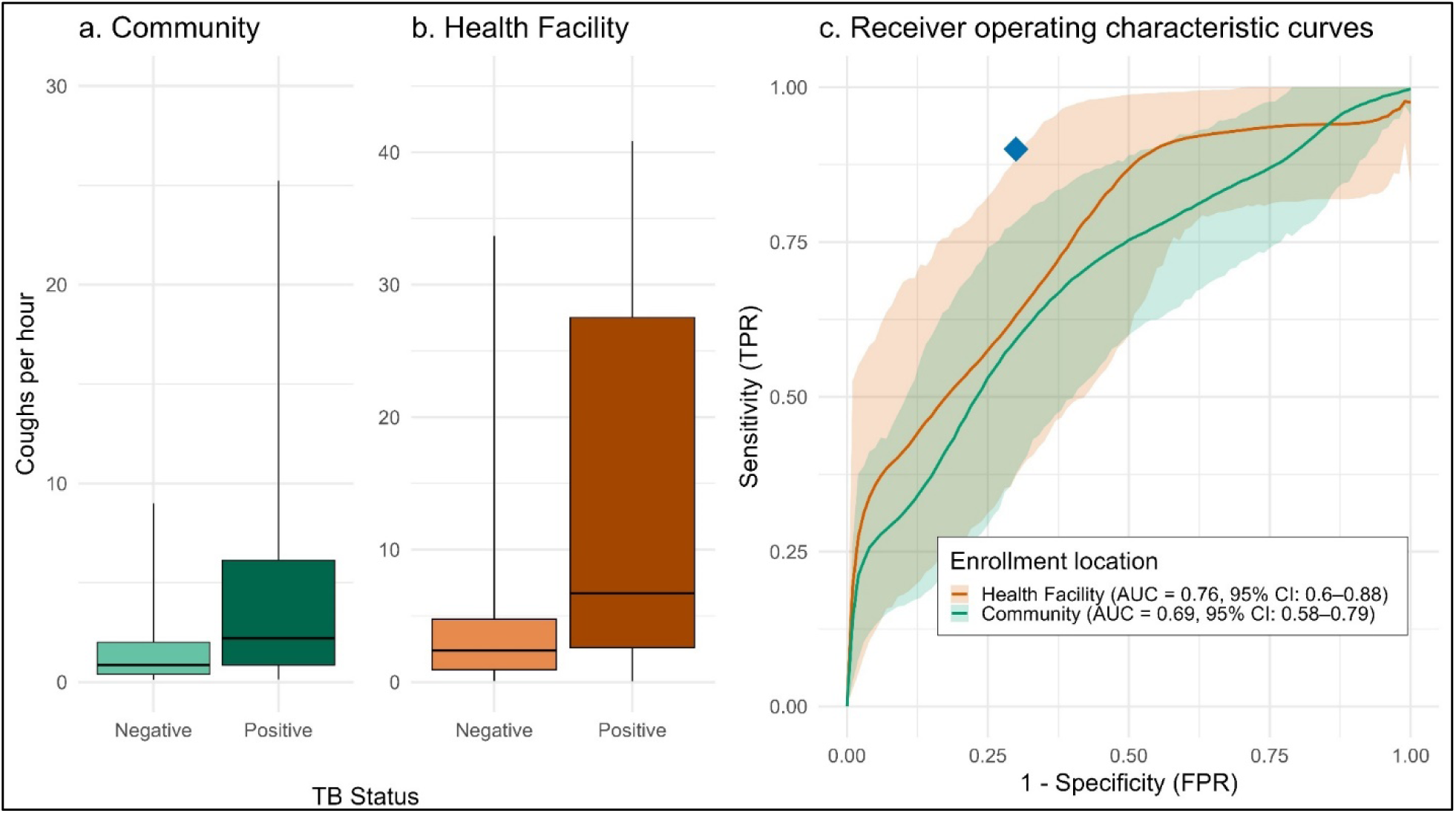
Relationship between weighted cough frequency and TB status, stratified by enrollment location. Boxplots show the distribution of measured cough frequency by TB status among community participants (panel A) and health facility participants (panel B). The central line within each box represents the median; the lower and upper edges of the box correspond to the 25th (Q1) and 75th (Q3) percentiles, respectively; and whiskers extend to the smallest and largest values. Panel C shows weighted receiver operating characteristic for cough frequency as a predictor for TB in both community (green) and health facility settings (orange). Shaded areas represent 95% confidence intervals, and the blue diamond represents the World Health Organization’s (WHO) 2014 target product profile for screening tests, of 90% sensitivity at a specificity of 70%.

At a threshold that would provide sensitivity of 90%, the specificity of self-reported cough as a diagnostic tool for TB was 20% (95% CI, 0%-22%) in the community cohort and 46% (95% CI, 1%-32%) in the health facility cohort. Conversely, at a specificity of 70%, sensitivity was 60% (95% CI, 37%-79%) in the community and 63% (95% CI, 37%-88%) in the health facility.

Recorded cough frequency had an AUC of 0.69 (95% CI, 0.58-0.79) in the community and 0.76 (95% CI, 0.6-0.88) in the health facility. (Figure 2c). This was lower than the AUC for self-reported cough in the community (0.83 [95% CI, 0.75-0.90]), but higher than in the health facility (0.63 [95% CI, 0.57-0.68]).

In the community setting, self-reported cough at screening among those who completed cough recording had a sensitivity for microbiologically confirmed TB of 44% (12/27, 95% CI 26-64%) and a specificity of 59% (17/29, 95% CI 39-76%). Comparing self-reported cough screening to recorded cough frequency in these participants, a recorded cough frequency cutoff of 1.51 coughs per hour had equivalent specificity (59%) to self-reported cough, but had a significantly higher (*P* = 0.04) sensitivity of 63% (17/27, 95% CI 43-80%).

Recorded cough frequency was positively correlated with several subjective and observed cough measures, with variation by enrollment location (Figure 3). Among participants enrolled from health facilities, recorded cough frequency was moderately correlated with self-reported cough severity (ρ = 0.56, p < 0.001), SGRQ score (ρ = 0.23, p = 0.046), observed coughs in 5 minutes (ρ = 0.33, p = 0.0014), and coughs observed by staff during the interview (ρ = 0.48, p < 0.001). Correlation with self-reported cough duration at enrollment was weaker and not statistically significant (ρ = 0.14, p = 0.21). In the community setting, stronger correlations were observed for cough duration at enrollment (ρ = 0.27, p = 0.039), cough severity (ρ = 0.27, p = 0.038), SGRQ score (ρ = 0.37, p = 0.033), coughs in 5 minutes (ρ = 0.28, p = 0.005), and interview-observed coughs (ρ = 0.43, p < 0.001). In contrast, the correlation between recorded cough frequency and self-reported cough duration at screening was not statistically significant (ρ = 0.18, p = 0.41).

**Figure 3.**
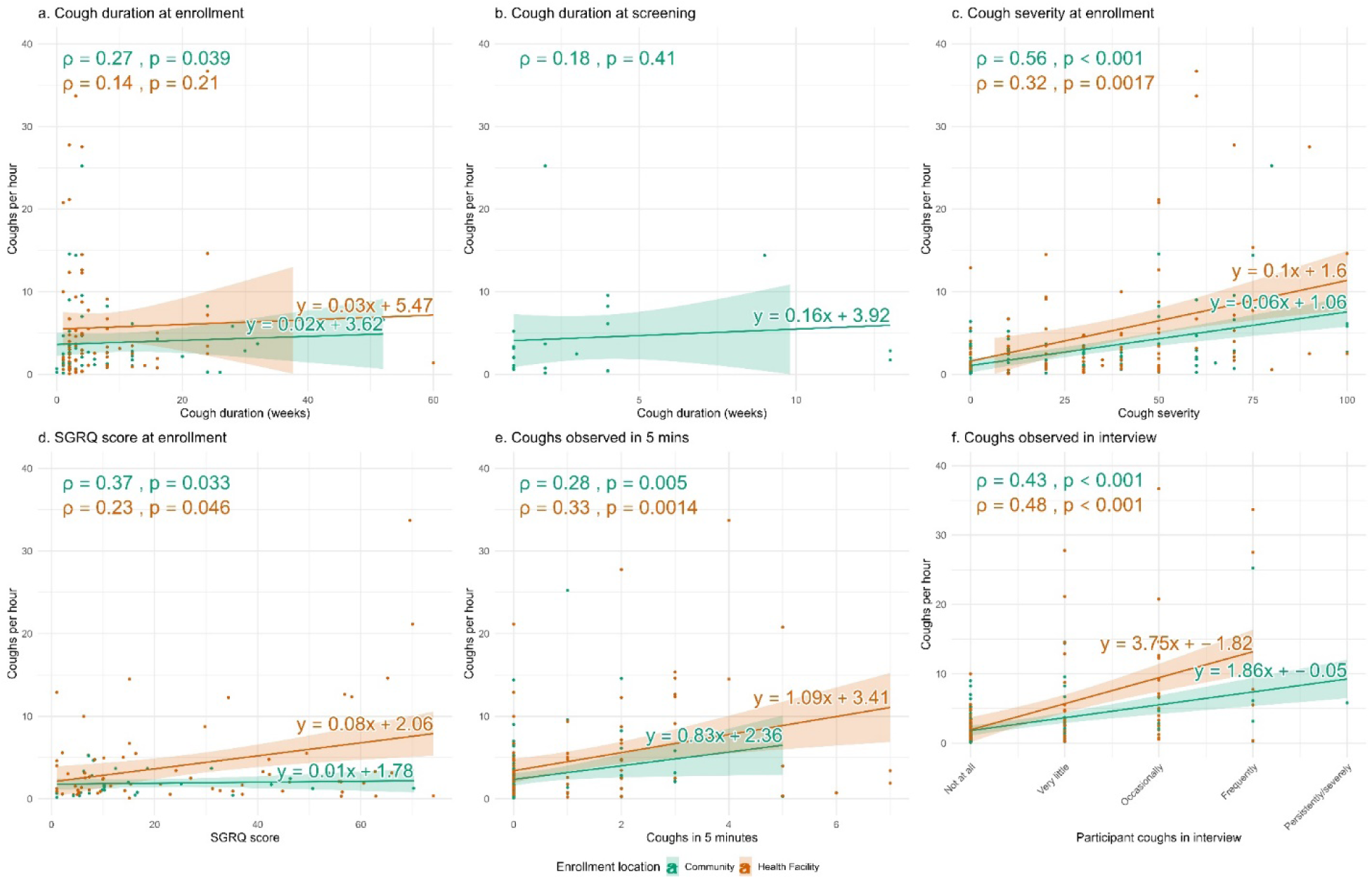
Associations between Hyfe app recorded cough frequency and participant-reported or observed indicators, stratified by enrollment location. Each plot includes estimated regression lines with 95% confidence intervals. Green points and lines represent community participants; orange represents health facility participants. Pearson correlation coefficients (ρ) and p-values are provided for each group. SGRQ: Saint George’s Respiratory Questionnaire

### Participant and staff feedback

Eight research staff were interviewed about their experience collecting cough-recording data. They described logistical and social barriers that resulted in low completion rates (Table 2). For staff, perceived security risks, unstable housing, alleged/untrustworthiness by community leaders, and unreliable contact information/frequent travel often drove decisions not to offer the smartphone-based recording device to participants. Staff noted that although the device was generally easy to set up, usually requiring 5 to 10 minutes, participants unfamiliar with smartphones often struggled with basic functions like charging from the provided power bank. Upon returning the device, many participants also shared that they had felt discomfort about the visibility of the microphone in public settings. Research assistants suggested several improvements to support broader use, including real-time monitoring features and more discreet, wearable designs with fewer visible components.

**Table 2.** Qualitative experience of using cough recording devices, as observed by research staff.

| Theme | Representative Quotes |
| --- | --- |
| The device was easy to use and set up | “Setting up the device can take like one minute but explaining the entire process... can take 5 to 8 minutes.”<br>“It is just an app that was put on a smartphone... it starts immediately to do the recording.”<br>“Setup is very easy... just having mobile data and logging into the Hyfe research app” |
| Participants had concerns about safety, stigma, and carrying the phone | “Some participants... were concerned people would think they were spying because of the microphone.”<br>“My boyfriend thought I had another man because of the phone; I had to return it.”<br>“Participants feared to go to the market or bar with the phone... it’s about security and privacy” |
| Participants saw value in the device but had mixed reactions | “Some said it was positive if the phone could give them feedback on their cough monitoring.” |
|  | “Some were excited that the device could help diagnose TB without sputum.”<br>“Participants saw it as improving their treatment or differentiating TB from normal cough” |
| Future improvements should focus on hardware, privacy, and RA support | “We need devices that don’t require microphones or internet.”<br>“Make a smaller device like an earpiece that can record discreetly.”<br>“Give RAs dashboard access so they can see data in real-time” |

## Discussion

This study aimed to assess the diagnostic performance and implementation feasibility of smartphone-based cough frequency recording for differentiating individuals with versus without TB in both community-based and health facility-based settings in Kampala, Uganda. While people with TB had significantly higher cough frequency than those without TB in both cohorts, cough frequency alone did not meet the WHO target product profile benchmarks for TB screening or triage tests (90% sensitivity and 70% specificity) [9]. Objectively measured cough frequency was only weakly correlated with subjectively reported cough presence, severity, and durations. In measuring cough frequency, staff encountered significant implementation challenges including participant concerns about device visibility and safety, which limited completion of cough recording. These findings suggest that, while recorded cough frequency may signal important TB disease-related differences and could add value as part of a broader diagnostic strategy, currently available recording apps are unlikely to be useful as standalone tests for TB.

Although cough frequency alone fell short of consensus targets, its association with TB status highlights its potential utility. Prior studies by Lee et al. and Turner et al. have demonstrated that objectively measured cough is a stronger predictor of TB disease severity than self-reported symptoms, with elevated cough linked to greater infectiousness [18,19]. These findings suggest that high cough frequency among individuals with TB may serve as an indicator of both clinical significance and transmission risk. Identifying individuals with elevated cough burdens, even in the absence of definitive diagnostic confirmation, is likely still essential for interrupting transmission. Diagnostic performance may also be enhanced with further integration of AI. AI-enabled tools that incorporate acoustic, spectral, and statistical features of cough sounds have shown promise in improving detection and classification of coughs [20–22]. Collectively, these findings support the potential role of smartphone-based cough monitoring as a complementary tool within multi-modal TB detection strategies. That said, both self-report and acoustic recording approaches are susceptible to errors that can affect the accuracy of cough assessment.

The weak correlation of measured cough frequency with reported cough symptoms may reflect differences in the dimensions of cough captured by each measure, although there are also factors that limit the accuracy of each. Individuals with chronic respiratory conditions, allergies, or smoking history may perceive their cough as less severe, viewing it as a typical part of their baseline health rather than a sign of illness. Factors such as age, sex, concern about underlying disease, recall bias, and cultural norms may also affect how symptoms are reported. Thus, objective measures such as app-based recorded cough frequency over 48 hours may offer greater reliability. However, recorded cough frequency also faces several potential sources of error.

Cough has been shown to vary from one day to the next, perhaps due to natural variation or to variability in activities and environmental allergen or irritant exposures. Our 48-hour cough monitoring period may therefore have been too short to accurately estimate each participant’s average cough frequency [23,24]. Furthermore, in this study’s crowded urban environment, ambient noises may have been misclassified as cough, or sounds of others coughing nearby may have been misattributed to participants. Although the use of lapel microphones aimed to improve the specificity of cough attribution, some measurement errors are likely to have remained.

Implementing longitudinal cough recording in our urban Ugandan research setting posed substantial logistical challenges, resulting in a low proportion of participants with complete and usable data. Barriers such as limited smartphone familiarity, concerns about device visibility, and unstable housing conditions reduced both participant interest and staff willingness to distribute recorders consistently. These barriers made it more difficult to conduct extended cough monitoring. Future efforts could benefit from making devices smaller and less conspicuous, for example with wrist-worn sensors or pendant-like microphones, and from designing with extended battery life to reduce dependence on regular access to electricity. In addition, efforts to integrate real-time feedback mechanisms, enhance ambient noise filtering, or incorporate hybrid sensor approaches (e.g., combining acoustic and location data to increase noise filtering in known crowded locations) could improve recording accuracy in busy environments.

Implementation in settings with higher levels of smartphone ownership and greater digital literacy may support longer recording periods and improve data completeness. Collectively, these adaptations could enhance the feasibility, acceptability and diagnostic value of cough monitoring as part of TB screening strategies.

Although this study offers novel insights on cough frequency in a TB screening context, there are some limitations to its accuracy estimates. First, participant sampling was uneven by initial Ultra results, requiring inverse probability weighting to more accurately reflect the underlying screening and diagnostic populations. Second, completion rates were low due to operational barriers. This may have introduced selection bias, and in particular, the discretion exercised by study staff in in in offering devices may have led to underrepresentation of individuals with social and environmental risk factors such as unstable housing or crowded living conditions.

Third, TB status classification was based on microbiological results from expectorated sputum, using a combination of Ultra (in those without known prior TB) and culture testing. Inadequate sensitivity of expectorated sputum could have caused some people with TB to be misclassified as having indeterminate or negative TB status. This misclassification may have been greatest among those with fewer respiratory symptoms. Lastly, the lower performance of self-reported cough among those who completed cough frequency monitoring suggests that individuals who agreed to cough monitoring may not be representative of the full population, introducing potential selection bias related to symptom experience.

## Conclusion

While smartphone-based cough monitoring alone does not currently meet diagnostic performance thresholds for TB screening, it may offer complementary information about respiratory pathology when integrated with existing risk factors and screening tools. Future research should prioritize improving device acceptability, completeness of collected data, and the development of analytical methods for interpreting cough patterns to enhance both the feasibility and diagnostic value in high-burden, resource-limited settings.

## Data Availability

The deidentified dataset used for this study and a data dictionary are being uploaded to a controlled access data archive, per requirements of the institutional review board that reviewed and approved this study. The upload process is under review and this preprint will be updated with a link to the data as soon as available. At this time, all data produced in the present study are available upon reasonable request to the authors.

## Acknowledgements

We would like to thank all the study staff at Walimu and Makerere University for their hard work and dedication. We appreciate the guidance of faculty at Johns Hopkins in manuscript writing. Lastly, our work would not be possible without those who participated in our study.

## Author contributions

**Conceptualization**: Patrick Biché, Francis Kayondo, Emily Kendall

**Data curation**: Patrick Biché, Annet Nalutaaya

**Formal analysis**: Patrick Biché

**Funding acquisition**: Emily Kendall, David Dowdy, Achilles Katamba

**Investigation**: Francis Kayondo, James Mukiibi

**Methodology**: Patrick Biché, Francis Kayondo, Emily Kendall

**Project administration**: Patrick Biché, Annet Nalutaaya, Mariam Nantale

**Supervision**: Emily Kendall, David Dowdy, Achilles Katamba

**Writing – original draft**: Patrick Biché

**Writing – review and editing**: Patrick Biché, Francis Kayondo, Annet Nalutaaya, James Mukiibi, Mariam Nantale, Joowhan Sung, David Dowdy, Achilles Katamba, Emily Kendall

## List of abbreviations

AI: Artificial intelligence
AUC: Area under the curve
CI: Confidence interval
CXR: Chest x-ray
HIV: Human immunodeficiency virus
IQR: Interquartile range
ROC: Receiver operating characteristic
SGRQ: Saint George’s Respiratory Questionnaire
TB: Tuberculosis
TPP: Target product profile
WHO: World Health Organization

## Supplementary Material

### S1 Appendix

Symptom questionnaire during enrollment (for health facility cohort)

1. Which of the following symptoms do you have currently? (Currently can mean today or within the past few days.)

Select all that apply:

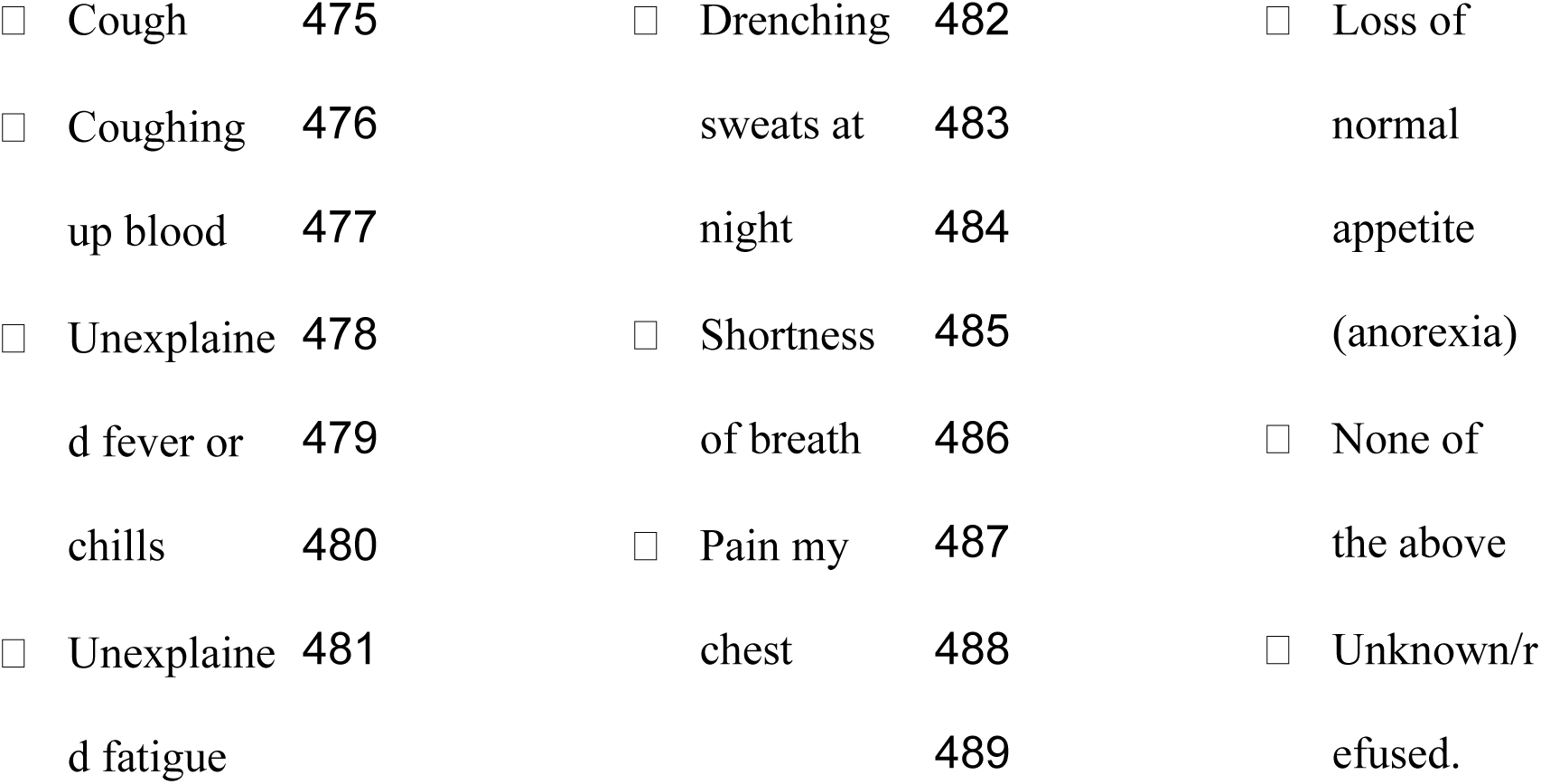

1. You said that you currently have a cough. Now looking back in time, for how long have you had this cough?

(Interviewer should record answer in weeks.)

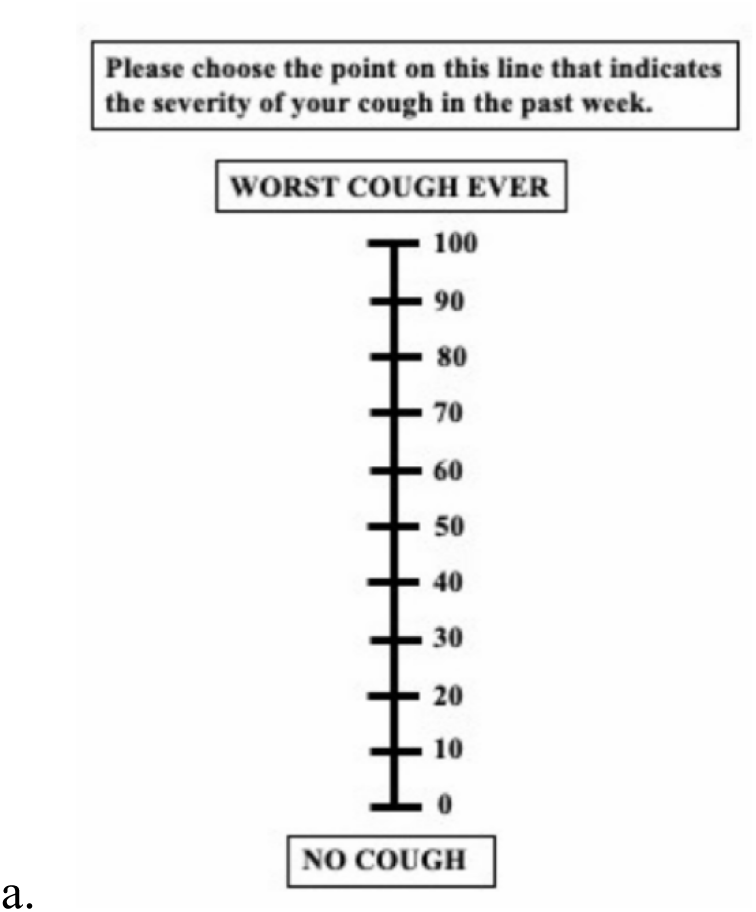

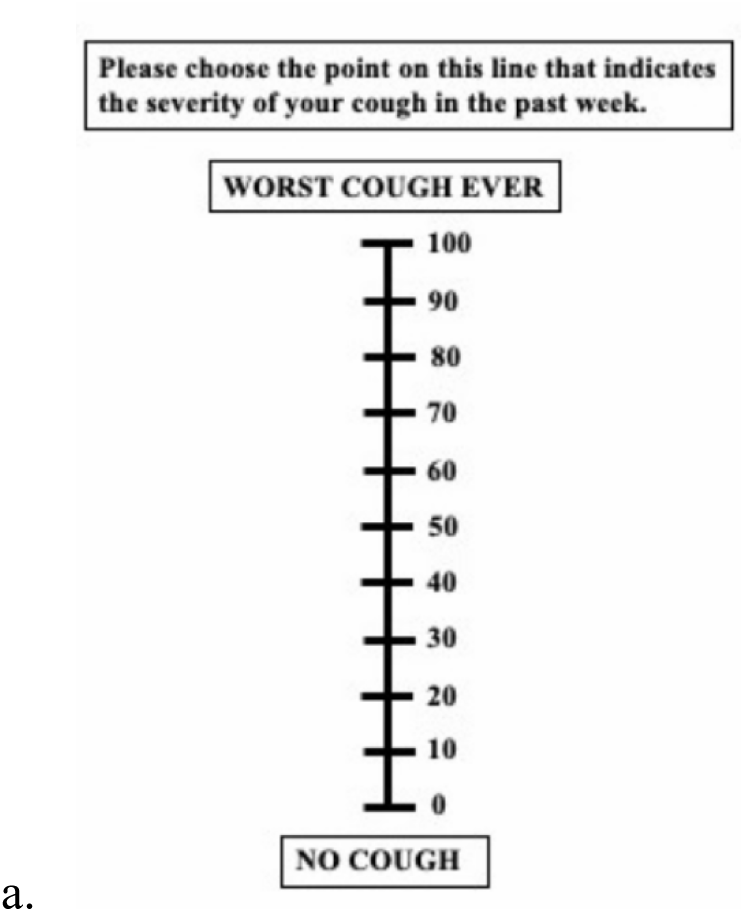

1. Use the visual scale below to indicate the severity of the cough in past week:
2. Number of coughs during a 5-minute observation period (as observed by person administering the CRF)
3. To be answered by the RA: How much did the participant cough during this interview?
4. Not at all
5. Very little (e.g. mild cough one or two times)
6. Occasionally (e.g. slight cough 3-5 times, or 1-2 moderate coughing episodes
7. Frequently (e.g. a frequent light cough, or 3-5 moderate coughing episodes”)
8. Persistently/severely.

Symptom questionnaire during screening (for community cohort)

1. TB sometimes has no symptoms, so we are testing everyone for TB regardless of whether or not they have symptoms. But we would like to know what symptoms you currently have. Do you have cough?
2. (If yes) For how many days have you had cough? Use the visual scale below to indicate the severity of the cough in past week:
3. Number of coughs during a 5-minute observation period (as observed by person administering the CRF)
4. To be answered by the RA: How much did the participant cough during this interview?
5. Not at all
6. Very little (e.g. mild cough one or two times)
7. Occasionally (e.g. slight cough 3-5 times, or 1-2 moderate coughing episodes
8. Frequently (e.g. a frequent light cough, or 3-5 moderate coughing episodes”)
9. Persistently/severely.

### S2 Appendix

Staff Interview Questions

1. Can you describe how you explained the cough recording device to participants?
2. How would you describe the ease of use (in respect to the setup) of the device?
3. How long (in minutes) did it usually take to set up and explain the device to participants?
4. Can you describe any negative feedback you received from participants?
5. Can you describe any positive feedback you received from participants?
6. What factors play into your decision not to give the device to a participant?
7. How can the device/software be improved for future studies?
8. How can future studies be improved to better support RAs/staff in implementing cough recorders?

9. Any other feedback you would like to share with the team?

**S3 Table.** Inverse probability weights.

| **Enrollment location** | **Xpert Ultra result** | **Proportion among those tested (%)** | **Proportion among those enrolled (%)** | **Weight** |
| --- | --- | --- | --- | --- |
| Community | Negative | 99.0 | 35.3 | 0.98 |
|  | Trace | 0.4 | 35.3 | 0.004 |
|  | Positive | 0.6 | 29.4 | 0.006 |
| Health Facility | Negative | 91.4 | 30.6 | 0.91 |
|  | Trace | 0.9 | 42.1 | 0.006 |
|  | Positive | 7.7 | 27.3 | 0.085 |

## Notes

### Competing Interest Statement

The authors have declared no competing interest.

### Funding Statement

This work was supported by grants from the National Institutes of Health (R01HL153611 [to E.A.K.], R01HL138728 [to D.W.D.], and K23AI185268 [to J.S.]) and the Gates Foundation (grant number INV-042921 to E.A.K.). The content is solely the responsibility of the authors and does not necessarily represent the official views of the funders.

## References

1. Frascella B, Richards AS, Sossen B, et al. Subclinical tuberculosis disease—a review and analysis of prevalence surveys to inform definitions, burden, associations, and screening methodology. Clin Infect Dis. 2021;73:e830–41.

2. Stuck L, Klinkenberg E, Abdelgadir Ali N, et al. Prevalence of subclinical pulmonary tuberculosis in adults in community settings: an individual participant data meta-analysis. Lancet Infect Dis. 2024;24:726–36.

3. Turner RD, Birring SS. Measuring cough: what really matters? J Thorac Dis. 2023;15:2288– 99.

4. Gabaldón-Figueira JC, Keen E, Giménez G, et al. Acoustic surveillance of cough for detecting respiratory disease using artificial intelligence. ERJ Open Res. 2022;8:00053–2022.

5. Gheisari M, Ghaderzadeh M, Li H, et al. Mobile apps for COVID-19 detection and diagnosis for future pandemic control: multidimensional systematic review. JMIR Mhealth Uhealth. 2024;12:e44406.

6. Hyfe Inc. Hyfe Research App. Hyfe. Accessed 2025 May 21. Available from: https://www.hyfe.com/research

7. Galvosas M, Gabaldón-Figueira JC, Keen EM, et al. Performance evaluation of the smartphone-based AI cough monitoring app - Hyfe Cough Tracker against solicited respiratory sounds. F1000Res. 2023;11:730.

8. Chaccour C, Sánchez-Olivieri I, Siegel S, et al. Validation and accuracy of the Hyfe cough monitoring system: a multicenter clinical study. Sci Rep. 2025;15:880.

9. World Health Organization. High-priority target product profiles for new tuberculosis diagnostics: report of a consensus meeting. Geneva: WHO; 2014.

10. Sung J, Nantale M, Nalutaaya A, et al. The long-term risk of tuberculosis among individuals with Xpert Ultra “trace” screening results: a longitudinal follow-up study. medRxiv [Preprint]. 2025 Mar 20. doi:10.1101/2025.03.20.25324205

11. Visek C, Dalmat RR, Nalutaaya A, et al. Prevalence and predictors of tuberculosis in adults and adolescents with sputum trace Ultra results in two high-burden clinical settings. 2025 May 18.

12. Jones PW, Quirk FH, Baveystock CM, et al. A self-complete measure of health status for chronic airflow limitation. The St. George’s Respiratory Questionnaire. Am Rev Respir Dis. 1992;145:1321–7.

13. Martin Nguyen A, Bacci ED, Vernon M, et al. Validation of a visual analog scale for assessing cough severity in patients with chronic cough. Ther Adv Respir Dis. 2021;15:17534666211049743.

14. QSR International. NVivo qualitative data analysis software. Version 14. QSR International; 2023. Available from: https://www.qsrinternational.com/nvivo-qualitative-data-analysis-software/home

15. Theron G, Venter R, Smith L, et al. False-positive Xpert MTB/RIF results in retested patients with previous tuberculosis: frequency, profile, and prospective clinical outcomes. J Clin Microbiol. 2018;56:e01696–17.

16. Hocking TD. WeightedROC: fast ROC curve estimation for weighted data. R package version 1.0.3; 2022. Available from: https://CRAN.R-project.org/package=WeightedROC

17. R Core Team. R: a language and environment for statistical computing. Version 4.3.1. R Foundation for Statistical Computing; 2023. Available from: https://www.r-project.org/

18. Lee GO, Comina G, Hernandez-Cordova G, et al. Cough dynamics in adults receiving tuberculosis treatment. PLoS One. 2020;15:e0231167.

19. Turner RD, Birring SS, Darmalingam M, et al. Daily cough frequency in tuberculosis and association with household infection. Int J Tuberc Lung Dis. 2018;22:863–70.

20. Miranda IDS, Diacon AH, Niesler TR. A comparative study of features for acoustic cough detection using deep architectures. Annu Int Conf IEEE Eng Med Biol Soc. 2019;2019:2601–5.

21. Pramono RX, Imtiaz SA, Rodriguez-Villegas E. A cough-based algorithm for automatic diagnosis of pertussis. PLoS One. 2016;11:e0162128.

22. Shati A, Hassan GM, Datta A. COVID-19 detection system: a comparative analysis of system performance based on acoustic features of cough audio signals. arXiv [Preprint]. 2024 Jun 19. doi:10.48550/arXiv.2309.04505

23. Chung KF, Chaccour C, Jover L, et al. Longitudinal cough frequency monitoring in persistent coughers: daily variability and predictability. Lung. 2024;202:561–8.

24. Tarlo SM, Altman KW, Oppenheimer J, et al. Occupational and environmental contributions to chronic cough in adults: CHEST Expert Panel report. Chest. 2016;150:894–907.

